# Persistence of pneumococcal carriage among older adults in the community despite COVID-19 mitigation measures

**DOI:** 10.1101/2022.06.28.22276654

**Authors:** Anne L. Wyllie, Sidiya Mbodj, Darani A. Thammavongsa, Maikel S. Hislop, Devyn Yolda-Carr, Pari Waghela, Maura Nakahata, Anne E. Watkins, Noel J. Vega, Anna York, Orchid M. Allicock, Geisa Wilkins, Andrea Ouyang, Laura Siqueiros, Yvette Strong, Kelly Anastasio, Ronika Alexander-Parrish, Adriano Arguedas, Bradford D. Gessner, Daniel M. Weinberger

**Affiliations:** Department of Epidemiology of Microbial Diseases, Yale School of Public Health, New Haven, CT 06510, USA; Yale Center for Clinical Investigation, New Haven, CT 06510, USA; Medical and Scientific Affairs, Pfizer Inc, 500 Arcola Rd, Collegeville, PA, 19426, USA

**Keywords:** pneumococcus, saliva, surveillance, carriage, pandemic

## Abstract

**Background:** Reported rates of invasive pneumococcal disease were markedly lower than normal during the 2020/2021 winter in the Northern Hemisphere, the first year after the start of the COVID-19 pandemic. However, little is known about rates of carriage of pneumococcus among adults during this period.

**Methods:** Between October 2020-August 2021, couples living in the Greater New Haven Area were enrolled if both individuals were aged 60 years and above and did not have any individuals under the age of 60 years living in the household. Saliva samples and questionnaires regarding social activities and contacts and medical history were obtained every 2 weeks for a period of 10 weeks. Following culture-enrichment, extracted DNA was tested using qPCR for pneumococcus-specific sequences *piaB* and *lytA*. Individuals were considered positive for pneumococcal carriage when Ct-values for *piaB* were less than 40.

**Results:** We collected 567 saliva samples from 95 individuals aged 60 years and above (47 household pairs and one singleton). Of those, 7.1% of samples tested positive for pneumococcus by either *piaB* only (n=6) or both *piaB* and *lytA* (n=34), representing 22/95 (23.2%) individuals and 16/48 (33.3%) households over the course of the 10-week study period. Study participants attended few social events during this period. However, many participants continued to have regular contact with children. Individuals who had regular contact with preschool and school aged children (i.e., 2-9 year olds) had a higher prevalence of carriage (15.9% vs 5.4%).

**Conclusions:** Despite COVID-19-related disruptions, a large proportion of older adults carried pneumococcus at least once during the 10-week study period. Prevalence was particularly high among those who had contact with school-aged children, but carriage was not limited to this group.

## INTRODUCTION

Mitigation measures that have been used to reduce the burden of COVID-19 have also had a profound effect on the incidence of disease caused by other pathogens. Major respiratory viruses, including influenza, respiratory syncytial virus, and human metapneumovirus, largely disappeared as causes of disease during the 2020-21 winter season in the Northern hemisphere [1–3]. Invasive pneumococcal disease (IPD) declined sharply in the spring of 2020 across all age groups and did not return to near regular levels until spring or summer of 2021 [4–7].

It was initially assumed that the reduction in the incidence of IPD was due to reduced transmission of the bacteria resulting from the implementation of non-pharmaceutical interventions. Pneumococcus is commonly carried in the upper respiratory tract of young children, however, the prevalence of pneumococcal carriage in children was near normal levels during 2020-2021 [3,8]. This demonstrates that children were still being exposed to and acquiring pneumococcus but not getting sick. Therefore, the decline in IPD observed in children might instead be related to the absence of infection by common respiratory viruses, which are thought to increase the risk of severe pneumococcal disease [3]. Additionally, some of the decline in IPD in children and adults could also be related to reduced healthcare seeking or changes in diagnostic practices during the pandemic [9].

While transmission of pneumococcus among children continued at high levels during the first year of the pandemic, social distancing and other mitigation measures could have reduced the amount of contact and transmission from children to adults. Children are a major source of exposure of pneumococcus for the adult population [10–12], which was most evident from the sharp drop in pneumococcal disease in adults that occurred following the introduction of pneumococcal conjugate vaccines in children [13]. Therefore, we expected that reductions in contact between children and adults might have occurred during the COVID-19 pandemic, leading to some of the reduction in IPD observed in adult populations. We evaluated carriage rates among adults ≥60 years of age living in the community in the United States during 2020-21 who were participating in an ongoing longitudinal carriage study. Detailed information about their activities and contacts, along with their carriage status, provides important insights into the potential drivers of pneumococcal epidemiology in older adults.

## METHODS

### Ethics

This study was approved by the Institutional Review Board at Yale School of Medicine (protocol ID. #2000026100). Demographic data and samples were only collected after the study participant had acknowledged that they had understood the study protocol and provided verbal- or written-informed consent. All participant information and samples were collected in association with anonymized study identifiers.

### Enrollment and eligibility

These data are drawn from the first sampling year of an ongoing study of pneumococcal carriage. The broader study is designed to quantify and detect rates of acquisition of pneumococcal carriage among older adults and the role of household transmission between cohabitating older adults. We recruited pairs of individuals residing in the community in the greater New Haven area who were both 60 years of age or older and who did not have anyone under the age of 60 years living in the household. If an individual had symptoms of respiratory illness at time of consenting or had received antibiotics or pneumococcal vaccination within the past four weeks, the enrollment of that household pair into the study was delayed by up to four weeks. There were no exclusion criteria based on underlying health status.

### Sample and data collection

Household pairs were sampled every 2 weeks, for a total of six visits covering 10 weeks. At each visit, the participants provided a self-collected saliva sample [14] into an empty 25 mL Eppendorf conical tube and answered questions about their social activities, doctors’ visits, recent contact with children, and any respiratory symptoms they were experiencing or had experienced in the two weeks prior (Appendix). Study participants left their samples outside their front door for contactless-collection and transport back to the laboratory at room temperature.

### Sample processing and pneumococcal detection

On arrival at the lab, 100 μl of raw saliva was plated on TSAII plates with 5% sheep’s blood and 10% gentamicin and grown overnight at 37°C with 5% CO_2_. Growth was harvested into 2100 μl of BHI supplemented with 10% glycerol and stored at -80°C until further processing. These samples were considered as culture-enriched for pneumococcus [15]. DNA was extracted from 200 μl of each culture-enriched sample using the MagMAX Viral/Pathogen Nucleic Acid Isolation kit (ThermoFisher Scientific) on the KingFisher Apex (ThermoFisher Scientific) with a modified protocol. Briefly, samples first underwent an extended digestion step, with 10 μl of proteinase K added to each sample and were incubated at 56°C for 10 minutes followed by heat inactivation of the proteinase K at 95°C for 10 minutes. Binding buffer and magnetic beads (25 μl) were added separately before proceeding with the KingFisher Apex extraction protocol which included an additional elution step, eluting extracted DNA into two plates of 50 μl of elution buffer. Purified DNA was tested by qPCR using primers and probes specific for two pneumococcal genes: *piaB* [16,17] and *lytA* [18]. The assays were carried out in 20 µl reaction volumes using SsoAdvanced Universal Probe Supermix (Biorad, USA), 2.5 µl of genomic DNA and primer/probe mixes at concentrations of 250 nM (Iowa Black quenchers) for *piaB* (1 μl per reaction) and *lytA* (1.2 μl per reaction). DNA of *S. pneumoniae* serotype 19F was included as a positive control in every run. Assays were run on a CFX96 Touch (Biorad) under the following conditions: 95°C for 3 minutes, followed by 45 cycles of 98°C for 15 seconds and 60°C for 30 seconds. Because many other *Streptococci* have the *lytA* gene, a sample was only considered to be positive for pneumococcus if it was positive for *piaB*. Samples were classified as positive with a *piaB* cycle threshold (Ct) value of <40 by RT-qPCR.

### Strain isolation and serotyping

Cultured-enriched saliva samples in which a higher concentration of pneumococcus was detected (<30 Ct by qPCR) were re-visited in an attempt to isolate pure pneumococcus. Samples were serially diluted 10-fold over a range of 10^−1^ to 10^−5^ in 1X PBS. The 10^−5^ and 10^−6^ serially diluted samples were plated (100 μl) onto plain blood agar plates. After overnight incubation, culture plates were visually screened for pneumococcal-like colonies. Each colony of pneumococcus-like morphology was streaked onto a plain blood agar plate and then inoculated into 50 µl of elution buffer (ThermoFisher Scientific) in a microcentrifuge tube. From each saliva sample, a total of 20 colonies were streaked onto one plain blood agar plate, with colonies pooled by 5 in each microcentrifuge tube. Pooled bacterial colonies were incubated for 10 minutes at 95°C on a heating block, then tested in qPCR for *piaB* and *lytA*. Streaked isolates from the pooled boilate samples that generated any signal <40 Ct for either *piaB* or *lytA* were individually tested for optochin susceptibility. Optochin susceptible colonies which tested positive for both *piaB* and *lytA* were then serotyped by latex agglutination (Statens Serum Institut) [19].

### Detection of SARS-CoV-2

All saliva samples were also tested for the presence of SARS-CoV-2 virus RNA using the extraction-free SalivaDirect assay [20]. Briefly, 50 µl of each sample was heated at 95°C for 5 minutes before being tested in RT-qPCR for SARS-CoV-2 [21].

### Statistical analysis

Differences in the frequency of categorical outcomes were compared using Fisher’s Exact test in the R Statistical Software v4.1.2.

## RESULTS

### Population characteristics

From November 2020 through June 2021, 95 individuals from 48 households were sampled and completed all 6 visits. One household was composed of a single individual who was enrolled into the study due to residing in a living facility for older adults. The mean age was 71 years (range 60-86; **Figure S1**). Of the 570 samples collected, three were not tested due to low collection volume (n=1) or a weather-related delay (two weeks) in transporting samples to the lab (n=2). Of the study participants, 72% were white, and 42% had a bachelor’s degree or higher. Eight individuals had a positive test for SARS-CoV-2 prior to enrollment in the study. None of the participants reported a positive test for SARS-CoV-2 while enrolled in the study nor tested positive for SARS-CoV-2 in any of the collected samples.

### Prevalence of pneumococcal carriage

Overall, 40/567 (7.1%) of samples tested positive for pneumococcus based on *piaB*, with 22/95 (23.2%) individuals colonized on at least one time point (**Figure S2**). Several individuals were colonized at multiple timepoints including two individuals (participants 3 and 41) who were colonized throughout the 10 weeks sampling period and a third who was colonized at five of the six time points (participant 33). In 6/48 (12.5%) households, both members were carriers, though not necessarily at the same time point. When samples were positive for both *piaB* and *lytA* (n=34), there was good concordance in the bacterial density (Ct value) (**Figure S3)**. In some samples, the concentration of *lytA* was higher than *piaB* (lower Ct), likely reflecting the presence of non-pneumococcal *Streptococci* in addition to pneumococcus (**Figure S3**). In addition to higher specificity, the sensitivity of the *piaB* assay was also slightly higher, resulting in some samples near the limit of detection that were positive for *piaB* and negative for *lytA* (**Figure S3)**.

To confirm the qPCR results, we used traditional culture-based methods, which are feasible when a high concentration of pneumococcus is detected [16]. From the 40 samples that tested positive for *piaB*, five isolates, all from the same individual (participant 41), were successfully isolated and identified as pneumococcus. Each of the isolates was optochin sensitive, and the presence of both *piaB* and *lytA* genes were confirmed by qPCR. All isolates were identified as serotype 15B/C by latex agglutination.

### Reported activities outside of the home

During this period, which included the first winter of the COVID-19 pandemic, participants continued to take part in some social activities outside the home. The most commonly reported activities were gathering with family (40%) and friends (29%). Just 6% of participants reported taking part in activities at a community center, and 8% participated in fitness activities. The prevalence of pneumococcal carriage was modestly higher among those who reported activities with family (13.5%) compared with those participating in other social activities or those reporting no social activity (8.1%).

### Prevalence is higher among those with contact with children

The prevalence of pneumococcal carriage was substantially higher among individuals who had contact with children (13.0% vs 3.5%, *p*=0.002) (**Figure 1, Table 2)**. Participants who reported recent contact with <5 year olds and 5-9 year olds had elevated prevalence (17.5%, *p*=0.001; 15.9%, *p*=0.007, respectively, **Table 2**). Prevalence was not notably higher among those reporting contact with children older than 10 years of age (10.4%, *p*=0.47). While the numbers are sparse, further subdividing the <5 year old population demonstrates progressively higher prevalence among those reporting contact with children <12m (11.9%), 12-23 months (13.3%) and 24-59 months (20.5%).

**Figure 1.**
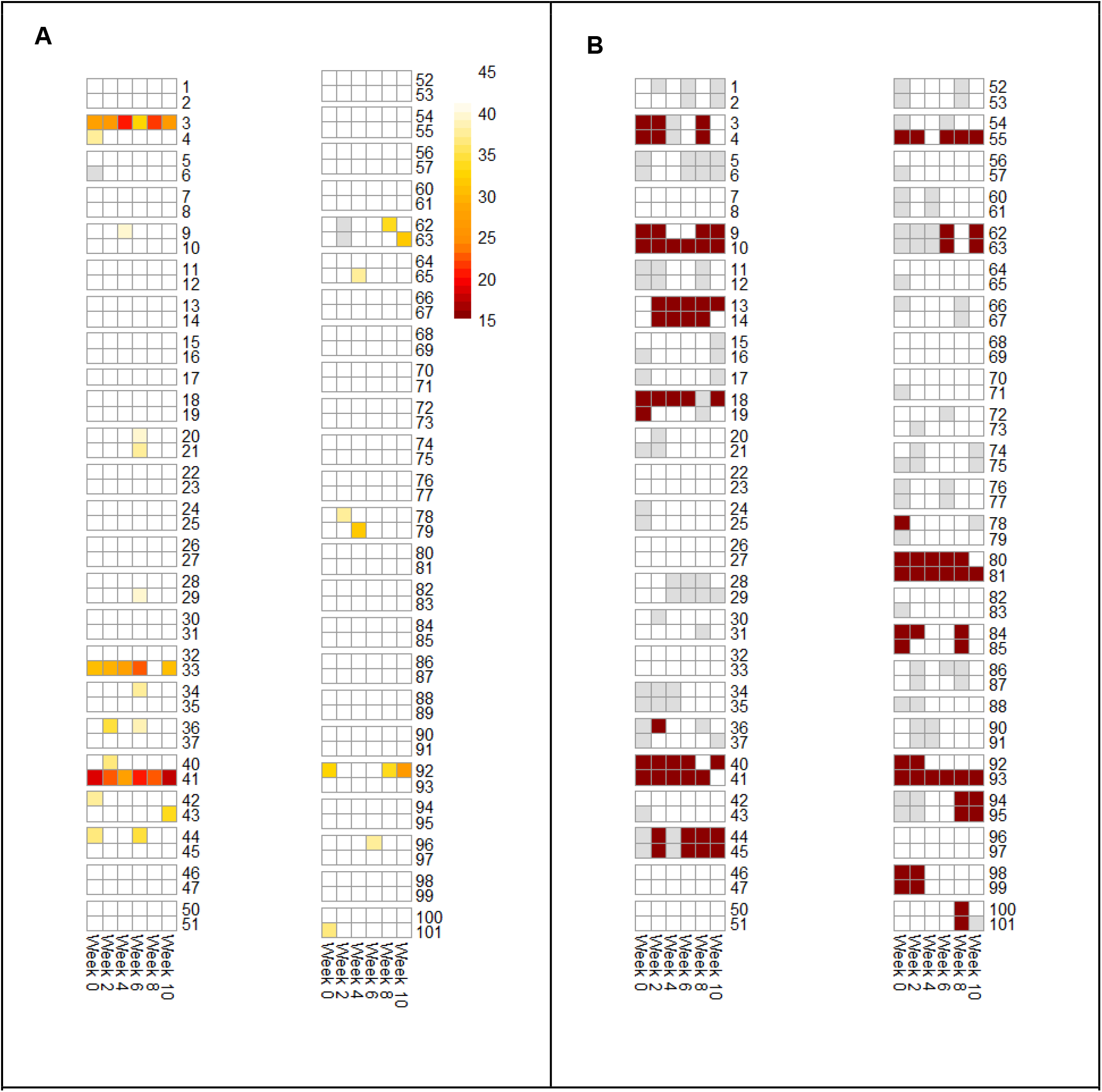
(A) Detection of pneumococcus as measured by Ct values from PCR assays targeting pneumococcal gene, *piaB*. Colored boxes indicate the individual is colonized, with darker colors indicating a higher presence of pneumococcus (lower Ct value). (B) Contact with children. Maroon indicates the individual reported contact with children, white indicates no reported contacts, gray indicates the question was not answered or the survey is missing.

**Table 1:** Relationship between pneumococcal detection and activities outside the home during the previous 2 weeks

|  | N* | Pneumococcus (*piaB*+) | Percent positive |
| --- | --- | --- | --- |
| Activities with family | 74 | 10 | 13.5% |
| Activities with friends | 38 | 0 | 0% |
| Fitness activities | 13 | 0 | 0% |
| Activities at community centers | 8 | 0 | 0% |
| Other social activity | 115 | 5 | 4.3% |
| Total* | 579 | 40 | 6.9% |

\*Numbers of visits at which people reported these activities. Participants were only asked about recent activities at visits 2-6.

**Table 2:** Relationship between pneumococcal detection and contact with children

|  | N | Pneumococcus (*piaB*+) | Percent positive |
| --- | --- | --- | --- |
| Contact with any children |  |  |  |
| No contact | 318 | 11 | 3.5% |
| Contact | 192 | 25 | 13.0% |
| Missing | 69 | 4 | 5.8% |
| Contact <5 year old children |  |  |  |
| No contact | 443 | 23 | 5.2% |
| Contact | 80 | 14 | 17.5% |
| Missing | 56 | 3 | 5.4% |
| Contact 5-10y old children |  |  |  |
| No contact | 441 | 24 | 5.4% |
| Contact | 82 | 13 | 15.9% |
| Missing | 56 | 3 | 5.4% |
| Contact >10y old children |  |  |  |
| No contact | 456 | 30 | 6.6% |
| Contact | 67 | 7 | 10.4% |
| Missing | 56 | 3 | 5.4% |

## DISCUSSION

Despite sharp declines in reported rates of IPD among adults during the first winter season of the COVID-19 pandemic (2020-21), older adults residing in the community continued to carry pneumococcus at levels consistent with what has been seen in other pre-pandemic studies of older adults that used similar molecular methods [15,22]. However, this study was conducted during a period when transmission mitigation measures related to the COVID-19 pandemic were in place. In the Greater New Haven Area, indoor mask mandates in public spaces were enforced and with high adherence [23], and usual community activities were canceled throughout the study period. Study participants reported few activities outside of the home. However, many individuals did continue to have regular contact with their families, including young children, and these participants had particularly high rates of carriage. This is consistent with studies conducted among younger adults that children are a major driver of transmission of pneumococcus in the community [24,25].

In this study setting, the period prevalence of pneumococcal carriage detected in this population of ≥60 year olds in the Greater New Haven Area was 23.2%. While other carriage studies conducted in older adults, also in the US, have reported lower rates of carriage [26], the difference in carriage rates can likely be attributed to sample and testing methodologies. As observed in the current study, saliva-based approaches and those that use qPCR following culture-enrichment tend to have higher sensitivity than those based on swabs and culture alone [15,16]. By including oropharyngeal swabs in addition to nasopharyngeal swabbing, Brache *et al*. reported a similar longitudinal carriage rate (≥25%), though did not detect an effect of contact with children [22]. In their analyses however, Branche *et al*. considered contact with any children under 5 years of age [22] and as our analyses show, children under 2 years of age contribute little to transmission [27]. This is also reflected in reports on their little contribution to disease incidence in older adults [27]. When contact with children is not stratified by age group, and the majority of child contacts are younger, the effect of transmission is harder to detect. In the current study, we asked study participants the ages of children with whom they interacted to enable these stratified analyses.

In this first study season of an ongoing study investigating rates of pneumococcal carriage in Greater New Haven, we found little evidence of impact of pandemic mitigation measures on rates of the carriage of pneumococcus in older adults. During this period of reduced social contact, study results suggest school-aged children are the likely source of continued presence of pneumococcus in most study subjects. Importantly, the high rate of non-specific signal detected in the widely used *lytA* qPCR assay demonstrates the importance of targeting multiple gene targets for reliable and specific detection of pneumococcus in oral samples. Follow-up studies with molecular serotyping of collected samples will provide greater insight into this observation and into transmission patterns of pneumococcus within households consisting of only individuals over the age of 60 years.

## Data Availability

All data produced in the present study are available upon reasonable request to the authors

## ACKNOWLEDGEMENTS

We thank the study participants for their time and dedication to our study.

## AUTHOR’S CONTRIBUTIONS

ALW, AA and DMW conceived the study. ALW, RAP, AA, BDG and DMW designed the study protocol. ALW, SM, YS, KA, MN and DMW managed the study. GW, AO, LS and YS collected the data. SM, DAT, MSH, DYC, PW, NJV, AY and OMA were responsible for sample receipt, processing and testing. ALW, DAT, MSH, MN, AEW and DMW performed the analyses and interpreted the data. ALW, DAT, MSH and DMW drafted the manuscript. All authors amended and commented on the final manuscript.

---

**Figure S1:**
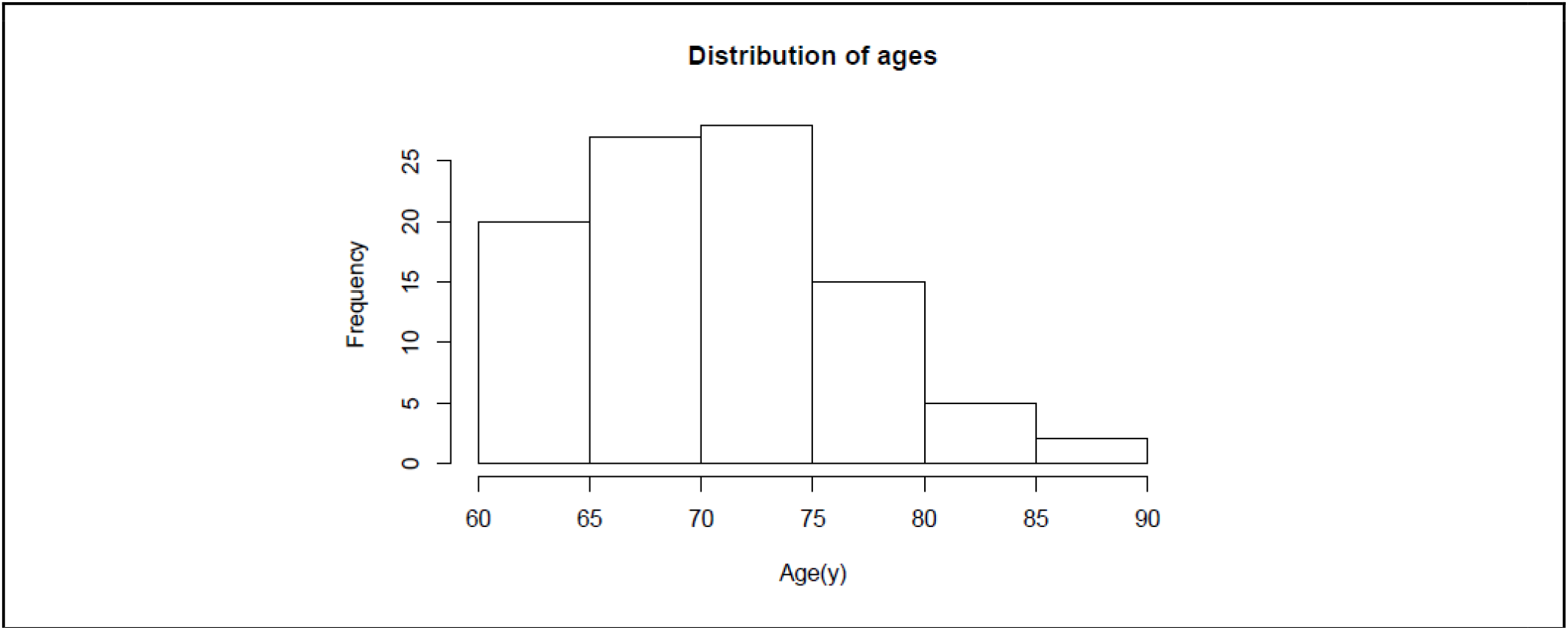
Age distribution of study participants.

**Figure S2.**
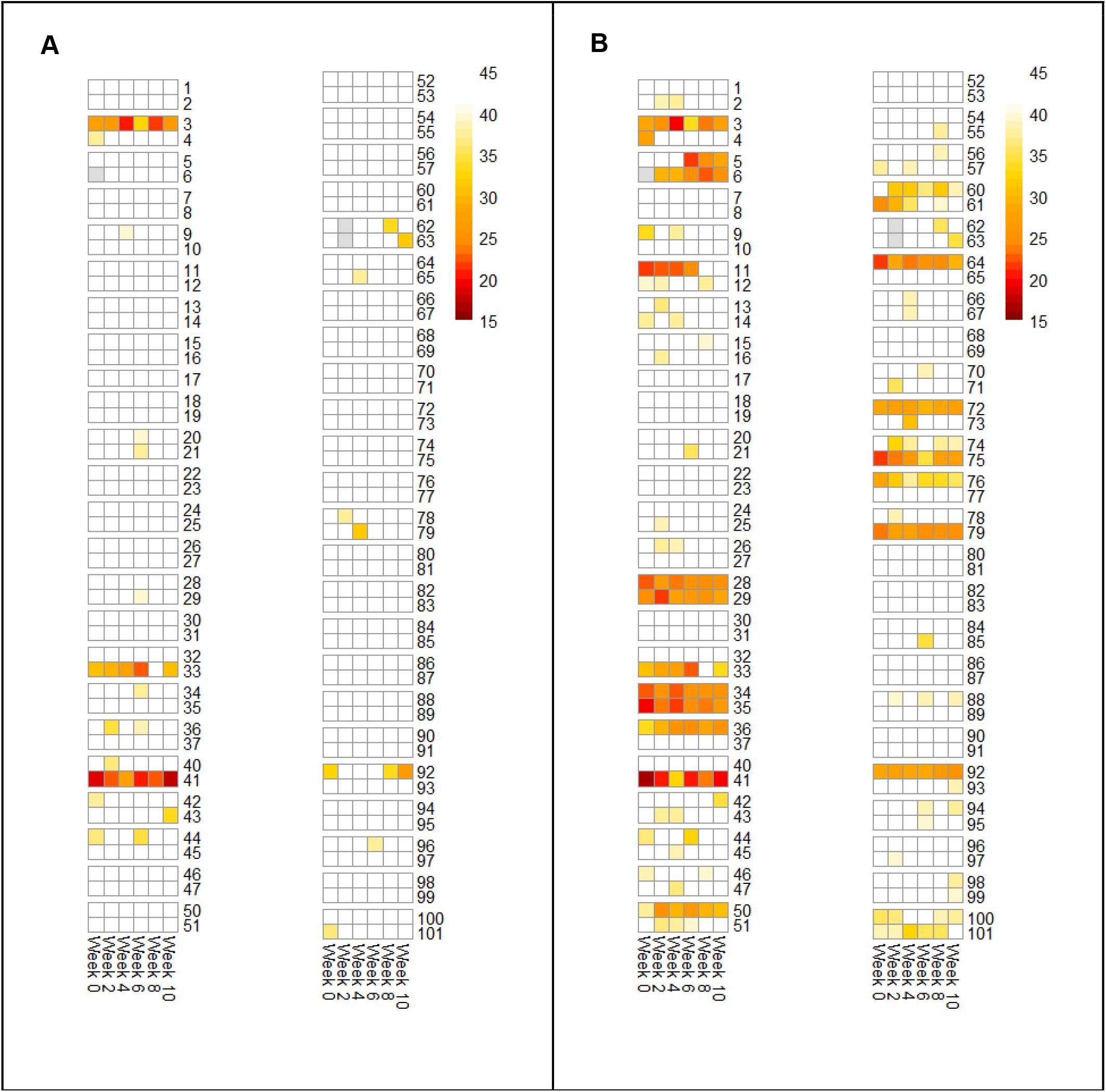
Detection of pneumococcal genes, (A) *piaB* and (B) *lytA* in saliva samples from household pairs aged 60 years and older, collected December 2020 through June 2021. Each row represents a study participant, and each column is a sample (each collected 2 weeks apart). Darker colors indicate higher density of bacteria (lower PCR Ct value), white indicates no detection of the gene target, gray indicates the sample was not tested.

**Figure S3.**
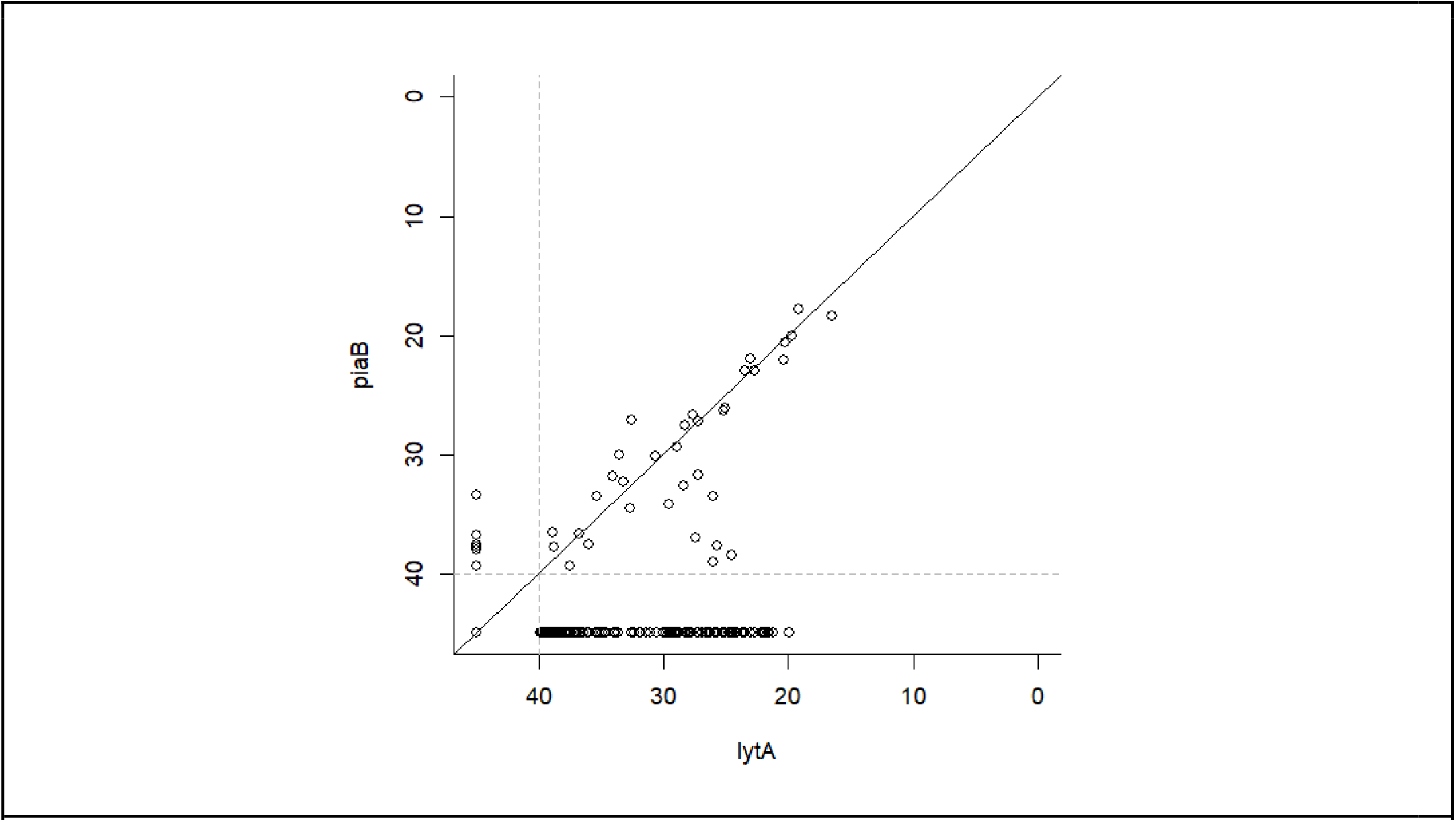
PCR Ct values for pneumococcus genes *piaB* and *lytA* detected in saliva samples from household pairs aged 60 years and older, collected December 2020 through June 2021. Values along the diagonal line indicate equal concentrations for both gene targets.

